# Hip geometric parameters are associated with radiographic and clinical hip osteoarthritis: findings from a cross-sectional study in UK Biobank

**DOI:** 10.1101/2023.03.28.23287740

**Authors:** SV Heppenstall, R Ebsim, FR Saunders, C Lindner, JS Gregory, RM Aspden, NC Harvey, T Cootes, JH Tobias, M Frysz, BG Faber

**Author notes:** Corresponding Author Dr Benjamin Faber, Musculoskeletal Research Unit, Learning and Research Building, Southmead Hospital, Bristol BS10 5FN. Joint senior author (M Frysz, BG Faber).

## Abstract

**Objectives:** To examine the extent to which geometric parameters derived from dual-energy x-ray absorptiometry (DXA) scans in the UK Biobank (UKB) study are related to hip osteoarthritis (HOA) independently of sex, age and body size.

**Methods:** Femoral neck width (FNW), diameter of the femoral head (DFH) and hip axis length (HAL) were derived automatically from left hip DXA scans in UKB using outline points placed around the hip by a machine-learning program. Correlations were calculated between geometric parameters, age, height, and weight. Logistic regression was used to examine the relationship of geometric parameters with radiographic hip osteoarthritis (rHOA), and hospital diagnosed HOA (HESOA), and Cox proportional hazards model to evaluate the relationship with total hip replacement (THR). Analyses were adjusted for sex, age, height, weight, and geometric parameters.

**Results:** Complete data were available for 40,312 participants. In age and sex-adjusted analyses, FNW, HAL and DFH were all related to increased risk of rHOA. Despite strong relationships between geometric parameters and body size, relationships between geometric parameters and HOA showed little attenuation after adjustment for height and weight. Following mutual adjustment, both HAL and FNW retained independent relationships with rHOA, while DFH was now protective. Only FNW was independently related to HESOA and THR.

**Conclusion:** Greater FNW and HAL were independently related to an increased risk of rHOA, whereas greater DFH appeared to be protective. Greater FNW was independently predictive of HESOA and THR. These results suggest DXA-derived geometric parameters, particularly FNW, could help to predict HOA and THR risk.

## Introduction

Osteoarthritis (OA) is a major cause of pain and disability globally, with the hip being the third most commonly affected joint (1). Morphological variation in hip shape has long been postulated as a risk factor for the development of hip OA (HOA) (2-5). To explore these associations, geometric parameters of hip shape have been measured on 2-dimensional imaging. Thus, femoral neck width (FNW), hip-axis length (HAL) and diameter of the femoral head (DFH) have been shown to associate with HOA when examined individually in small studies, but the inter-relatedness of these measures have not been explored previously (6, 7).

Hip shape is known to vary greatly between the sexes with females having a larger neck shaft angle and smaller femoral head and neck (8, 9). However, geometric parameters measuring femur size are intrinsically related to body size and each other (6). This has made understanding independent associations between geometric parameters and HOA difficult especially given previous studies have tended to examine aspects of shape in isolation without sex stratification (6, 7). Statistical Shape Modelling (SSM) has evolved as an alternative approach to quantifying hip morphology and captures the whole of the hip joint. This method does overcome the issue of geometric parameters being correlated with size and each other, however the main limitation of SSM is that it is challenging to determine which specific aspects of hip shape are related to the outcome of interest. Therefore, geometric parameters can provide complementary information, hence why we have decided to look at them separately.

The availability of large cohorts with hip dual-energy X-ray absorptiometry (DXA) scans linked to HOA outcomes, such as in the UK Biobank (UKB) study, provides an excellent opportunity to examine relationships between geometric parameters and HOA in more detail. Improvements in scan resolution have shown images acquired with newer DXA scanners to be suitable for ascertaining both hip shape and radiographic HOA (rHOA) measures (10, 11). In addition, DXA scans involve lower doses of radiation than traditional radiographs, offering the potential for use in screening. Existing methods such as hip structural analysis (HSA) are already available for deriving geometric parameters such as FNW and HAL for hip DXA scans (12), however this does not generate other parameters potentially related to HOA such as DFH.

The UKB study has now undertaken ∼40,000 high resolution hip DXA scans. This large sample now offers opportunities to explore relationships between geometric parameters and HOA as defined both clinically and radiographically. To understand the relationship between geometric parameters and HOA, in the present study, we aimed to: (i) determine the correlation between geometric parameters and measures of body size, (ii) describe the cross-sectional relationships between geometric parameters with rHOA and hospital diagnosed hip OA (HESOA), (iii) describe the longitudinal associations between geometric parameters and total hip replacement (THR) and (iv) establish which of these are independent as assessed in mutually adjusted models.

## Materials and Methods

### Population

UKB is a large prospective study, which at baseline (2006-2010), recruited over 500,000 men and women aged 40-69 years (13). Participants have undergone extensive phenotypic assessments through questionnaires, imaging, physical measures, and electronic healthcare record linkage (14). In 2014 UKB commenced the extended imaging study with the aim of conducting DXA scans on 100,000 participants. DXA scans were obtained from both hips (iDXA GE-Lunar, Madison, WI), with participant’s limbs being positioned with 15-25° internal rotation using a standardised protocol. UKB has ethical approval from the National Information Governance Board for Health and Social Care and North-West Multi-centre Research Ethics Committee (11/NW/0382) which covers this study (application number 17295). All participants gave informed written consent.

### Dual-energy X-ray absorptiometry variables

#### Outline points and radiographic osteoarthritis annotation

A machine-learning algorithm placed 85 points to outline the proximal femur and acetabulum in all available left hip DXA scans as of April 2021 (11). Each image was checked, and points were corrected if necessary (∼90% of images required no correction) and at the same time osteophytes were manually annotated using a custom tool (The University of Manchester) (Figure 1). The outline points did not encompass any annotated osteophytes. rHOA grades 0-4 were assigned semi-automatically to each hip DXA image combining osteophyte and joint space width data as previously described (10).

**Figure 1:**
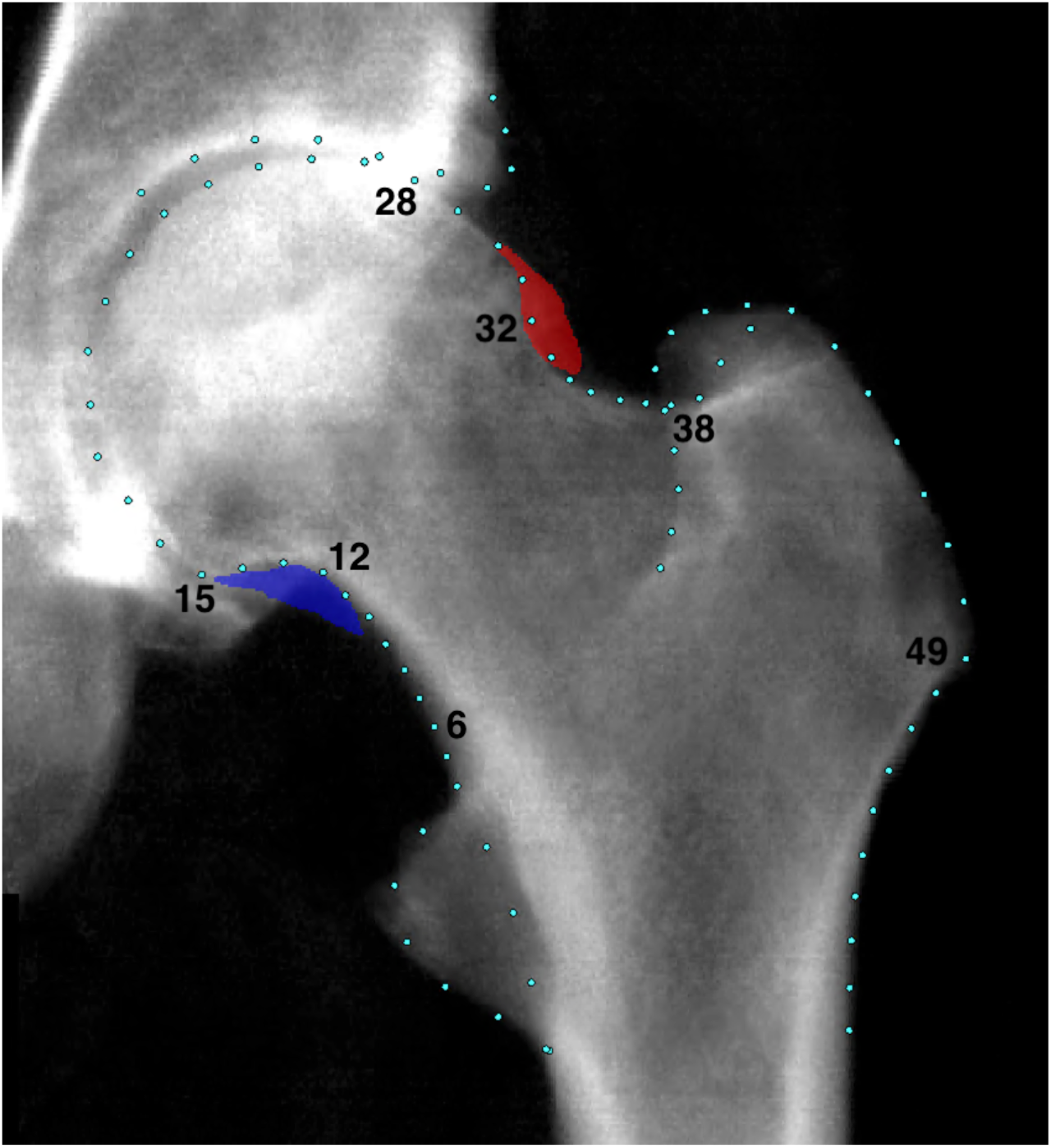
An example of a grade 4 rHOA hip from UK Biobank with superolateral (red) and inferomedial (blue) osteophytes marked.

#### Geometric parameters

Custom Python 3.0 scripts were developed and used to automatically derive FNW, DFH and HAL. These scripts are openly available (15). The DXA DICOM images store pixel dimension data facilitating the calculation of geometric parameters in millimetres (mm). FNW was defined as the shortest distance measured between the superior and inferior side of the femoral neck (16). To measure this, points 6-12 defined the inferior side and points 32-38 defined the superior side of the femoral neck (Figure 1). A line-segment approach was used to automatically calculate the narrowest distance between these points (Figure 2A), a description of this approach has been published previously (17). DFH was defined as the distance across the spherical aspect of the femoral head. To estimate this, a circle of best fit was placed around the femoral head using a least-squares package in Python that was applied to points 15-28 (18). The diameter of the circle was taken to represent the DFH in millimeters (Figure 2B). HAL was defined as the distance from the base of the greater trochanter to the medial aspect of the femoral head in millimeters. In previous studies, HAL measured using HSA software included medial joint space width as it is measured to the inner pelvic rim (19). In this study however, outline points were not reliably available for the medial acetabulum hence the measure only encompassed the femur. To measure this, a straight line was drawn from point 49 through the centre of the circle of best fit (used to calculated DFH). HAL was calculated from point 49 to where the line intersected the circumference of the circle after it has passed through the centre point of the circle (Figure 2C). All images with values +/- 2 standard deviations (SDs) from the mean were reviewed manually. In addition, FNW and HAL measures derived automatically were compared with values derived from HSA software (iDXA GE-Lunar, Madison, WI) in a subset of participants to check comparability.

**Figure 2:**
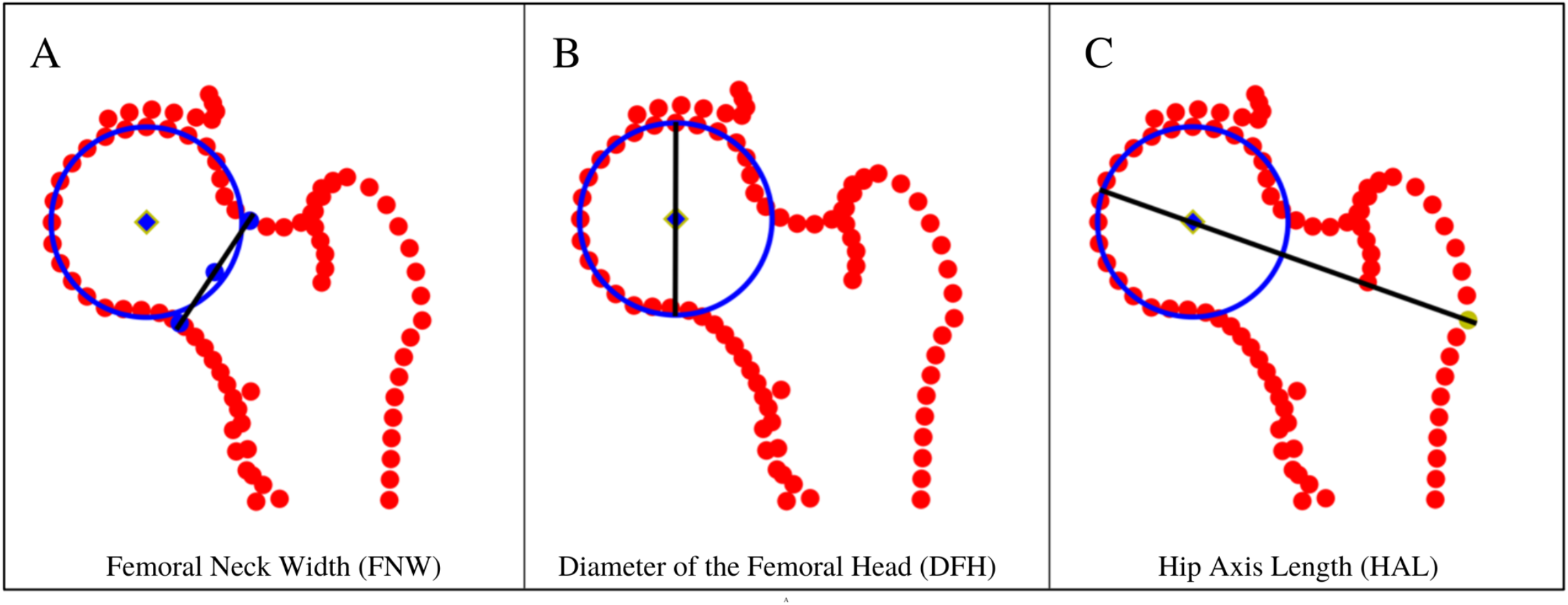
Geometric parameter measurements: A – Femoral Neck Width (FNW) derived using the line-segment method using points 6-12 on the medial side and points 32-38 on the lateral side, B- Diameter of the femoral head (DFH) derived by fitting a circle to points 15-28, C – Hip Axis Length (HAL) derived by finding the distance between point 49 and where the line intersects the circumference of the circle having passed through the centre of the femoral head.

### Clinical outcomes

HESOA data were obtained from hospital episode statistics (HES). THR data were based on the Office of Population Censuses and Surveys Classification of Interventions and Procedures codes (ICD-10) obtained from HES data. Further information on how these data were derived has been described previously (10). The clinical outcomes used in this study were not side-specific.

### Statistical analysis

Baseline characteristics are shown as means, SDs and ranges for continuous variables and as frequencies for categorical variables. Correlations between geometric parameters, height, weight, and age were derived using Pearson’s correlation test statistic (r^2^). Distributions of continuous variables were checked visually for normality. Frequencies were used for categorical variables. Correlation values ranging between r^2^ ≥ 0.7-1.0, r^2^ ≥ 0.5-0.7, and r^2^ < 0.5 were deemed as strong, moderate and weak correlations respectively. As we anticipated these to be correlated, a sensitivity analysis was done where the ratio of DFH to FNW was used as an exposure and compared with the associations between DFH, and clinical and radiographic outcomes in models 1-4. Logistic regression was used to examine associations between each standardised exposure (FNW, DFH and HAL) and each HOA outcome (grade ≥2 rHOA and HESOA). Sensitivity analyses were carried out using grade ≥3 rHOA and grade 4 rHOA only as outcomes. Results are presented as odds ratios (OR) with 95% confidence intervals (CIs) and p values. Cox proportional hazard modelling was used to investigate the associations between standardised geometric parameters and THR and results are presented as hazard ratios (HR) with 95% CIs and p values. The Cox proportional hazard assumptions were tested and met. We present unadjusted (model 1), and confounder adjusted analyses (partially adjusted = models 2, 3 and fully adjusted = model 4). Potential confounders were defined *a priori*, and these were included in partially adjusted models as covariates (model 2: adjusted for age and sex; model 3: model 2 plus height and weight to account for effects of body size with which geometric parameters are known to be strongly related) (6, 20-22). A final model also included mutual adjustment for other geometric parameters (fully adjusted/model 4). Sex-stratified analyses were conducted due to known differences in hip shape. All statistical analyses were performed using STATA version 17 (Stata Corp, College Station, TX, USA). Furthermore, a glossary of terms is provided for the reader in Supplementary Table 1.

## Results

### Baseline characteristics

A total of 40,312 individuals (mean age 63.7, SD 7.6 years, range 44-82) had left hip DXAs available for analysis (Table 1). 21,021 (52.1%) participants were women and 19,291 (47.9%) were men. rHOA grade ≥2 was present in 3,014 (7.5%) individuals, rHOA grade ≥3 in 700 (1.7%) and rHOA grade 4 in 157 (0.4%). Males had a higher prevalence of rHOA across all three grades compared with females. 527 (1.3%) individuals had HESOA and 259 (0.6%) underwent THR after their hip DXA. In contrast with rHOA, the prevalence of clinical outcomes was higher in females compared with males (Table 1).

**Table 1:**
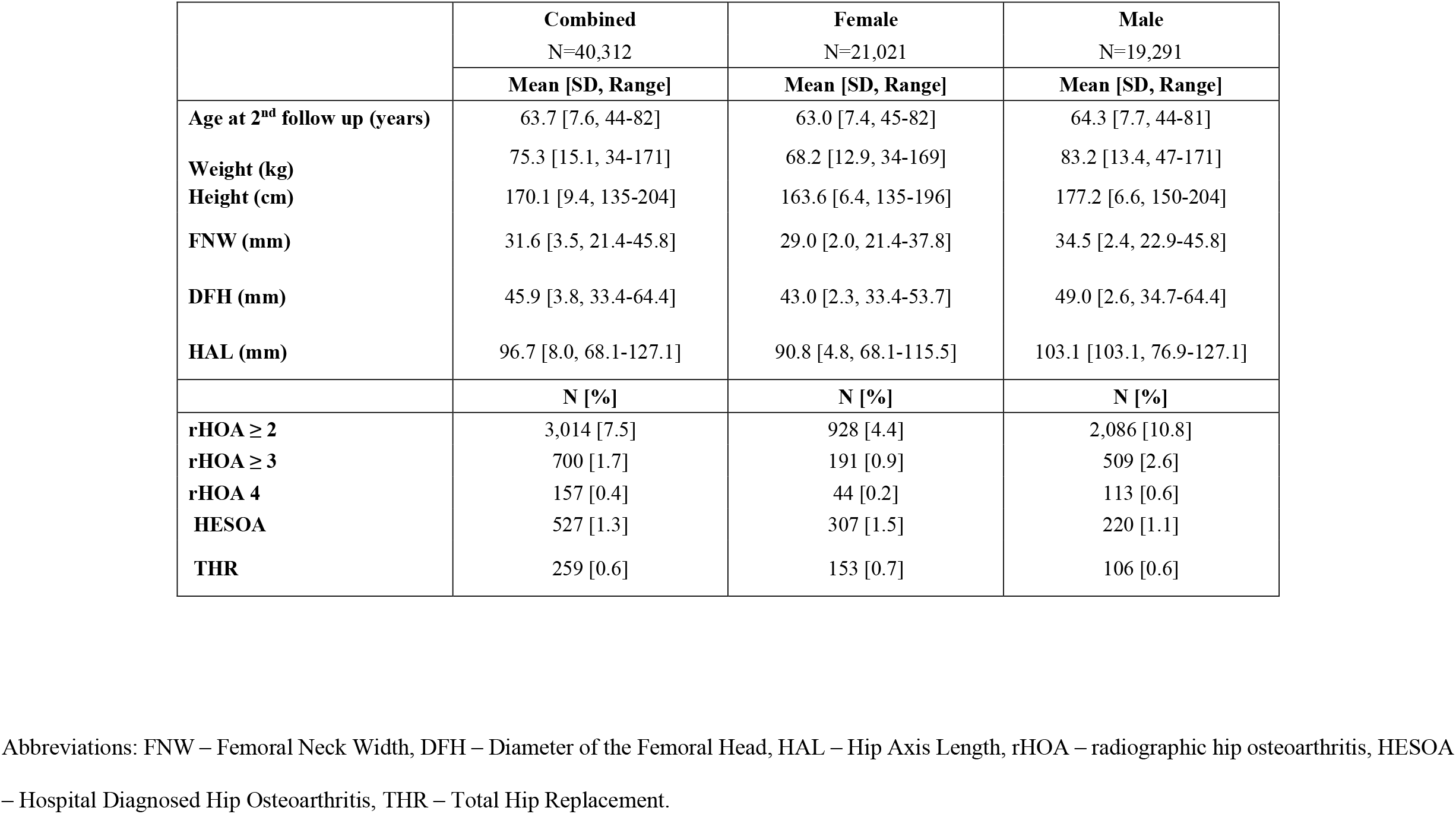
Population Characteristics

### Geometric parameters and their inter-relationships

The mean FNW was 31.6 mm (SD 3.5, range 21.4-45.8), DFH 45.9 mm (3.8, 33.4-64.4) and HAL 96.7 mm (8.0, 68.1-127.1). Males had a greater FNW, DFH and HAL compared with females [FNW: male mean 34.5 mm (SD 2.4, range 22.9-45.8) / female mean 29.0 mm (SD 2.0, range 21.4-37.8), DFH: 49.0 mm (2.6, 34.7-64.4)/ 43.0 (2.3, 33.4-53.7) & HAL: 103.1 mm (5.5 76.9-127.1)/ 90.8 mm (4.8, 68.1-115.5)] (Table 1). Comparison with HSA derived FNW and HAL showed strong correlations between these measures (r^2^ 0.97 & 0.93 respectively), however the mean values derived with HSA were larger (Supplementary Table 2). FNW, DFH, HAL and height were all strongly correlated (r^2^ 0.75-0.89) (Table 2). Weight also showed moderate correlations with FNW, DFH, HAL and height (r^2^ 0.52-0.57).

**Table 2:** Correlation (r^2^) matrix between geometric parameters, height, weight, and age.

|  | **FNW** | **DFH** | **HAL** | **Height** | **Weight** | **Age** |
| --- | --- | --- | --- | --- | --- | --- |
| **FNW** | 1 |  |  |  |  |  |
| **DFH** | 0.89 | 1 |  |  |  |  |
| **HAL** | 0.82 | 0.87 | 1 |  |  |  |
| **Height** | 0.75 | 0.81 | 0.81 | 1 |  |  |
| **Weight** | 0.55 | 0.52 | 0.53 | 0.57 | 1 |  |
| **Age** | 0.13 | 0.11 | 0.13 | -0.06 | -0.07 | 1 |

Green indicates a strong correlation ( $r^2 \geq 0.7$ -1.0), yellow a moderate correlation ( $r^2 \geq 0.5$ - 0.7) and amber/red a weak correlation ( $r^2 < 0.5$ ). Abbreviations: FNW
– Femoral Neck Width, DFH – Diameter of the Femoral Head, HAL – Hip Axis Length.

### Femoral neck width versus HOA

In analyses adjusted for age, sex, height and weight (model 3) progressive associations between a wider FNW and higher rHOA grades were seen [rHOA grades ≥2: OR 1.81 (95% CI 1.70-1.94), grades ≥3: 3.52 (3.09-4.00) and grade 4: 5.11 (3.95-6.60)]. Similar results were also seen in unadjusted (model 1) and age and sex adjusted (model 2) analyses (Supplementary Table 3). If anything, associations with rHOA were strengthened when adjusting for sex, height and weight as well as geometric parameters (model 4) [grade ≥2 rHOA: OR 2.38 (95% CI 2.18-2.59), grade ≥3 rHOA: 5.26 (4.45-6.22) and grade 4 rHOA: 8.55 (6.11-11.95)] (Figure 3, Table 3). Further sex stratified analyses showed similar associations in females and males albeit female effect estimates tended to have wider confidence intervals (Supplementary Figure 1, Supplementary Tables 4 and 5). Partially adjusted sex combined analyses (model 3) showed an association between FNW and HESOA [OR 2.00 (95% CI 1.71-2.34)] and THR [HR 2.27 (95% CI 1.82-2.83)]. These associations were strengthened after adjusting for other geometric parameters [model 4: HESOA: OR 2.20 (95% CI 1.80-2.68), THR: HR 2.51 (95% CI 1.89-3.32)]. (Figure 3, Table 3).

**Figure 3:**
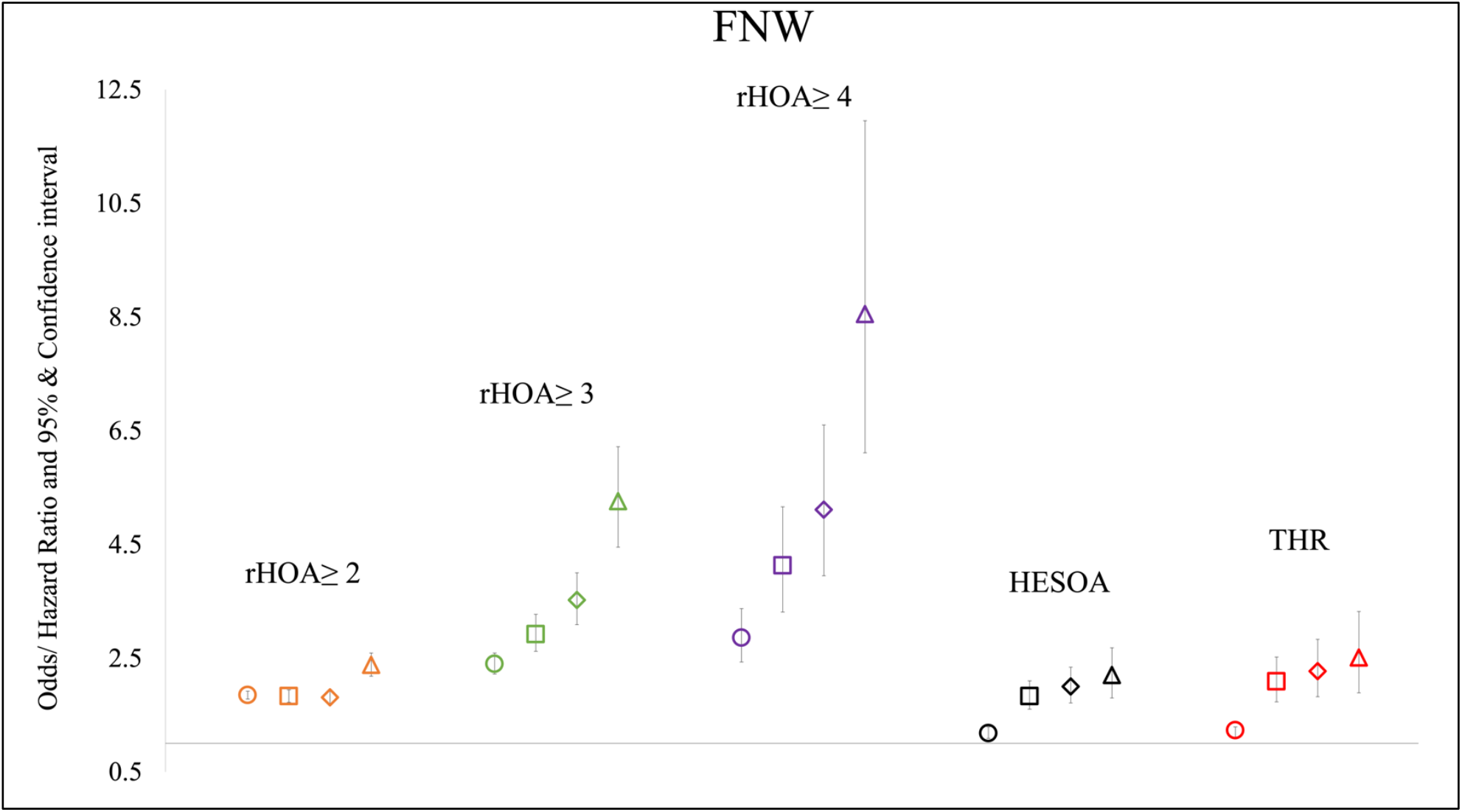
Logistic regression results for the associations between femoral neck width (FNW) and radiographic hip osteoarthritis ≥ grade 2, ≥ grade 3, grade 4 and hospital diagnosed OA (HESOA) and cox proportional hazard modelling results between FNW and total hip replacement (THR) in combined sex analyses. Each symbol represents odds/ hazard ratios with 95% CIs. Circle symbol represents unadjusted analyses (model 1), square indicates adjustment for age and sex (model 2), diamond for age, sex, height and weight (model 3) and triangle for age, sex, height, weight, and remaining geometric parameters (model 4).

**Table 3:** Logistic regression/ Cox proportional hazard modelling results showing the association between geometric parameters and HOA outcomes.

| | | Grade $\geq 2$ rHOA | | Grade $\geq 3$ rHOA | | Grade 4 rHOA | | HESOA | | THR | |
| --- | --- | --- | --- | --- | --- | --- | --- | --- | --- | --- | --- |
|  |  | OR [95% CI] | p | OR [95% CI] | p | OR [95% CI] | p | OR [95% CI] | p | HR [95% CI] | p |
| Model 3 | FNW | 1.81 [1.70-1.94] | $9.20 \times 10^{-69}$ | 3.52 [3.09-4.00] | $7.50 \times 10^{-82}$ | 5.11 [3.95-6.60] | $1.64 \times 10^{-35}$ | 2.00 [1.71-2.34] | $3.78 \times 10^{-18}$ | 2.27 [1.82-2.83] | $2.80 \times 10^{-13}$ |
| Model 4 | FNW | 2.38 [2.18-2.59] | $1.40 \times 10^{-88}$ | 5.26 [4.45-6.22] | $1.20 \times 10^{-84}$ | 8.55 [6.11-11.95] | $4.59 \times 10^{-36}$ | 2.20 [1.80-2.68] | $1.06 \times 10^{-14}$ | 2.51 [1.89-3.32] | $1.44 \times 10^{-10}$ |
| Model 3 | HAL | 1.28 [1.19-1.37] | $1.17 \times 10^{-11}$ | 1.64 [1.42-1.88] | $3.72 \times 10^{-12}$ | 1.66 [1.24-2.21] | $5.88 \times 10^{-4}$ | 1.29 [1.09-1.53] | $2.71 \times 10^{-3}$ | 1.49 [1.18-1.89] | $8.32 \times 10^{-4}$ |
| Model 4 | HAL | 1.25 [1.15-1.36] | $5.99 \times 10^{-8}$ | 1.39 [1.18-1.63] | $4.95 \times 10^{-5}$ | 1.28 [0.93-1.78] | 0.13 | 1.06 [0.88-1.29] | 0.52 | 1.23 [0.94-1.61] | 0.13 |
| Model 3 | DFH | 1.08 [1.01-1.16] | 0.03 | 1.49 [1.30-1.71] | $2.01 \times 10^{-8}$ | 1.73 [1.30-2.30] | $1.65 \times 10^{-4}$ | 1.41 [1.19-1.67] | $5.38 \times 10^{-5}$ | 1.50 [1.19-1.90] | $7.12 \times 10^{-4}$ |
| Model 4 | DFH | 0.55 [0.50-0.61] | $3.32 \times 10^{-32}$ | 0.42 [0.35-0.51] | $1.07 \times 10^{-17}$ | 0.36 [0.24-0.54] | $9.55 \times 10^{-7}$ | 0.82 [0.65-1.03] | 0.09 | 0.75 [0.54-1.04] | 0.09 |

Table shows logistic regression results for the associations between geometric parameters and radiographic hip osteoarthritis (rHOA) ≥ grade 2, ≥ grade 3, grade 4 and hospital diagnosed hip osteoarthritis (HESOA) and Cox proportional hazard modelling results between geometric parameters and total hip replacement (THR) in partially and fully adjusted combined sex analyses (model 3 included the covariates age, sex, height and weight whereas model 4 was adjusted for these as well as the remaining geometric parameters).

### Hip axis length versus HOA

In partially adjusted (model 3) sex combined analyses, there were associations between a longer HAL and higher rHOA grades [rHOA grades ≥2: OR 1.28 (95% CI 1.19-1.37), grades ≥3: 1.64 (1.42-1.88) and grade 4: 1.66 (1.24-2.21)]. These findings were consistent with unadjusted (model 1) and age and sex adjusted (model 2) analyses (Supplementary Table 3). On complete adjustment (model 4) there was some attenuation of effect between HAL and each rHOA grade [rHOA grades ≥2: OR 1.25 (95% CI 1.15-1.36), grades ≥3: 1.39 (1.18-1.63) and grade 4: 1.28 (0.93-1.78)] (Figure 4, Table 3). Sex stratified analyses showed stronger associations in females compared with males especially for the higher rHOA grades [model 4: female/male rHOA grade 4 OR 2.05 (95% CI 1.08-3.89) / 1.08 (0.74-1.57)] (Supplementary Figure 1, Supplementary Tables 4 and 5).

**Figure 4:**
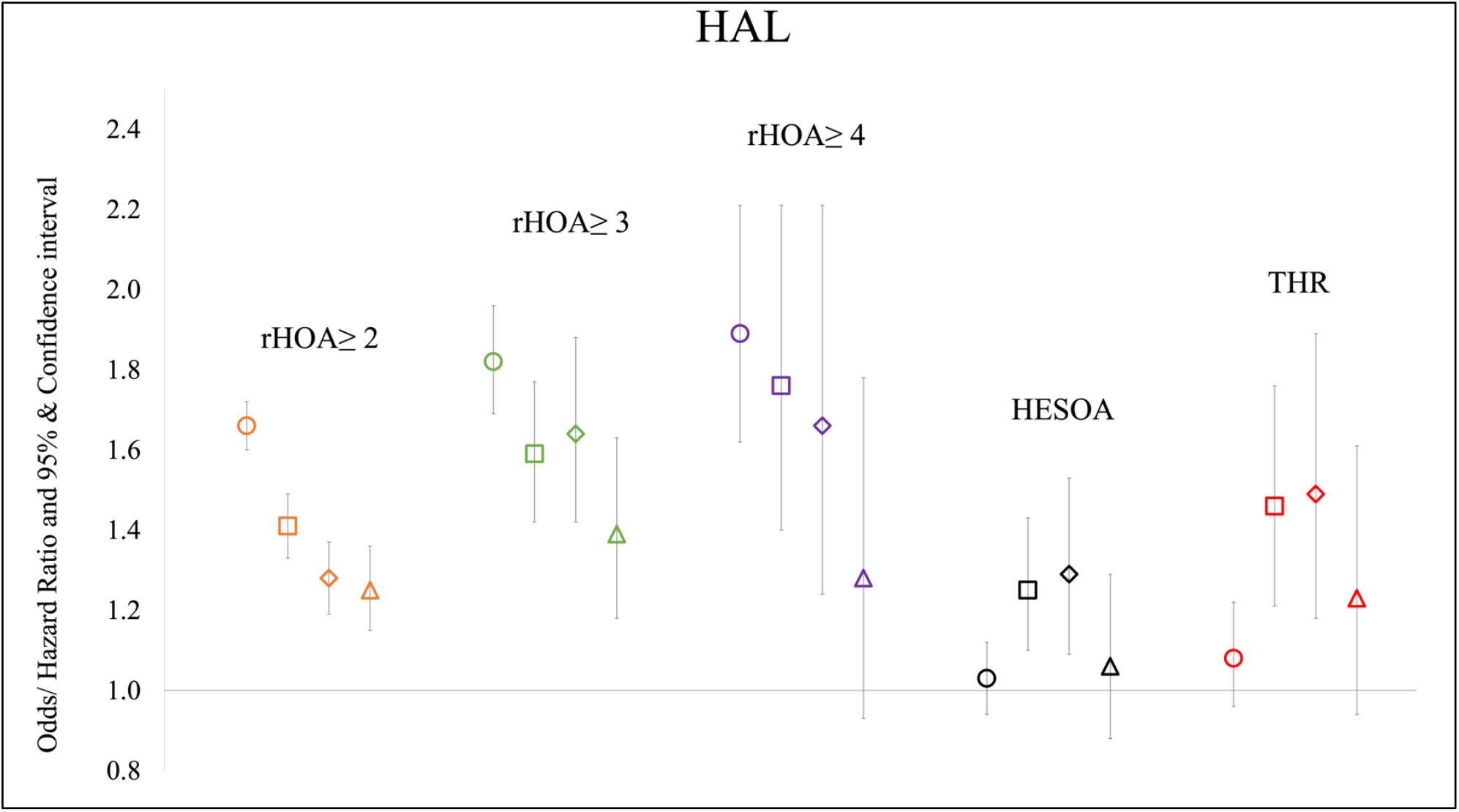
Logistic regression results for the associations between hip axis length (HAL) and radiographic hip osteoarthritis ≥ grade 2, ≥ grade 3, grade 4 and hospital diagnosed OA (HESOA) and cox proportional hazard modelling results between HAL and total hip replacement (THR) in combined sex analyses. Each symbol represents odds/ hazard ratios with 95% CIs. Circle symbol represents unadjusted analyses (model 1), square indicates adjustment for age and sex (model 2), diamond for age, sex, height and weight (model 3) and triangle for age, sex, height, weight, and remaining geometric parameters (model 4).

In partially adjusted (model 3) sex combined analyses there was some evidence of an association between HAL and HESOA and THR [HESOA: OR 1.29 (95% CI 1.09-1.53), THR: HR 1.49 (95% CI 1.18-1.89)]. After adjusting for other geometric parameters (model 4) these effect sizes were fully attenuated [HESOA: OR 1.06 (95% CI 0.88-1.29), THR: HR 1.23 (95% CI 0.94-1.61)]. (Figure 4, Table 3). Associations between HAL and HESOA and THR were broadly similar in the sex-stratified analyses (Supplementary Figures 2 and 3, Supplementary Tables 4 and 5).

### Diameter of the femoral head versus HOA

An association between a greater DFH and higher rHOA grades was present in partially adjusted (model 3) sex combined analyses [rHOA grade ≥2: OR 1.08 (95% CI 1.01-1.16), grade ≥3: 1.49 (1.30-1.71) and grade 4 rHOA: 1.73 (1.30-2.30)]. These effect sizes were reduced compared with the unadjusted (model 1) and age and sex adjusted (model 2) analyses (Supplementary Table 3). After adjusting for other geometric parameters, the direction of effect was reversed with increasing DFH displaying a protective effect with rHOA [rHOA grades ≥2: OR 0.55 (95% CI 0.50-0.61), grades ≥3: 0.42 (0.35-0.51) and grade 4: 0.36 (0.24-0.54)] (Figure 5, Table 3). Sex stratified analyses revealed similar results in females and males (Supplementary Figure 1, Supplementary Tables 4 and 5).

**Figure 5:**
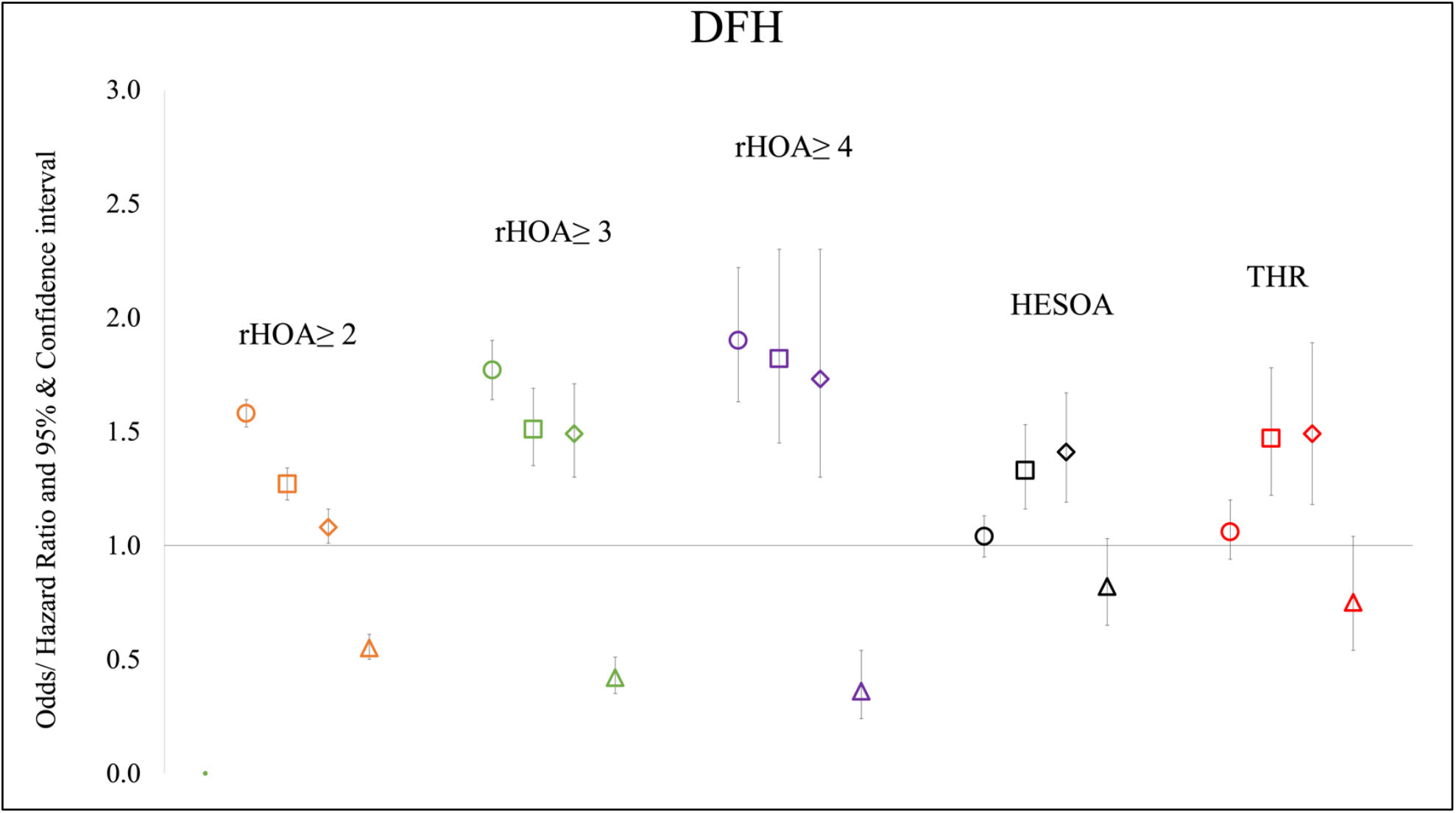
Logistic regression results for the associations between diameter of the femoral head (DFH) and radiographic hip osteoarthritis ≥ grade 2, ≥ grade 3, grade 4 and hospital diagnosed OA (HESOA) and cox proportional hazard modelling results between DFH and total hip replacement (THR) in combined sex analyses. Each symbol represents odds/ hazard ratios with 95% CIs. Circle symbol represents unadjusted analyses (model 1), square indicates adjustment for age and sex (model 2), diamond for age, sex, height and weight (model 3) and triangle for age, sex, height, weight, and remaining geometric parameters (model 4).

On partial adjustment (model 3) there was some evidence of an association of DFH with HESOA [OR 1.41 (95% CI 1.19-1.67)] and THR [OR 1.50 (95% CI 1.19-1.90)]. After adjusting for other geometric parameters (model 4) these associations attenuated [HESOA OR 0.82 (0.65-1.03)] and THR [HR 0.75 (95% CI 0.54-1.04)] (Figure 5, Table 3). Sex stratified results were similar to combined results in terms of magnitude and direction of effect (Supplementary Figures 2 and 3, Supplementary Tables 4 and 5).

Given the strong correlation between DFH and other geometric parameters, particularly FNW (Table 2), sensitivity analyses were conducted to confirm that the reversal of the association between DFH and rHOA following adjustment for other geometric parameters was not a spurious result due to collinearity between these variables. Therefore, we also examined whether DFH/FNW ratio showed equivalent relationships with HOA outcomes compared with DFH in our fully adjusted model (model 4). A similar inverse association between DFH/FNW and rHOA risk was observed in model 3 compared to that seen for DFH in model 4 (Supplementary Table 6). Moreover, the association between DFH/FNW and rHOA risk was unaffected by additional adjustment for geometric parameters. DFH/FNW was also associated with a reduced risk of other HOA outcomes namely HESOA and THR.

## Discussion

This is the largest observational study to date (N=40,312) exploring the associations between hip geometric parameters (FNW, DFH and HAL) and HOA. As expected, the geometric parameters were highly correlated with height and each other, as well as differing between the sexes. FNW, HAL and DFH were all related to increased risk of rHOA in age and sex adjusted analyses (model 2). Despite strong relationships between geometric parameters and height and weight, relationships between geometric parameters and HOA showed little attenuation once adjusted for height and weight. Following adjustment for geometric parameters (model 4) both HAL and FNW retained independent relationships with rHOA, whereas DFH was now protective. When considering the clinical outcomes, only FNW was independently related to HESOA and THR. These analyses suggest that a wider FNW, longer HAL and smaller DFH were associated with higher risk of rHOA. In contrast, only a wider FNW appeared to be associated with increased risk of HESOA and predictive of THR.

FNW showed the strongest associations with all HOA outcomes, consistent with findings from previous studies (6, 7, 23). In addition, these associations were strengthened when adjusted for body size (i.e., height and weight) and other geometric parameters which suggests that FNW is the leading geometric parameter associated with HOA, something which has not been shown before. Previous explanations for the association of FNW with HOA have focused on the mechanism of impingement of the widened femoral neck on the acetabulum akin to femoroacetabular impingement (23-25). An alternative explanation is that the femoral neck is widened as part of, or in parallel to the onset of HOA rather than the widened femoral neck causing HOA. Recent evidence from studies using SSM suggests a wider femoral neck is associated with more severe HOA and separately a genetic study suggested that a genetic predisposition to HOA led to cam-like changes at the femoral head (11, 26, 27). Both these studies suggest that hip shape is associated with but might not be a cause of HOA which has treatment implications.

DFH showed positive associations with HOA when considered in isolation, similar to a previous study (6), but when adjusted for the other geometric parameters the direction of effect reversed to show a relatively smaller femoral head was in fact a risk factor for rHOA. Equivalent findings were obtained after analysing relationships between DFH/FNW ratio and rHOA, and in addition, reduced DFH/FNW was a risk factor for HESOA and THR. A smaller femoral head has been shown to be associated with HOA before when measured on plain radiographs, with the authors suggesting a smaller contact area at the joint would raise the force going through it leading to degeneration (23). That said, these results are in contrast to previous studies based on SSM which reported associations with HOA to have both a larger femoral head and neck (11, 28, 29). This might be explained by how DFH is defined; for example, with a spherical hip, the diameter across the femoral head is the same when measured on multiple axes. Whereas, on an aspherical (cam-type) femoral head the elliptical nature means that the transverse diameter is greater than the vertical. In this study, the circle of best fit was fitted to the medial aspect of the femoral head so as not to be distorted by cam-type femoral heads. This is not the case for the aforementioned SSM studies. The corollary of this might be that a smaller spherical femoral head is a risk factor for HOA alongside larger aspherical (cam-type) femoral heads.

Whilst there was evidence of an association between HAL and HOA outcomes, these associations were fully attenuated following adjustment. Previously, when assessed in isolation a study had shown associations with HOA similar to model 3 in this study i.e. without adjustment for other geometric parameters (6). These results highlight the importance of considering measures of hip morphology alongside each other when examining relationships with HOA, as well as body size. Height and weight are both known risk factors for HOA and were correlated with the geometric parameters featured in this study (30).

In this study we have used a new method based on SSM points to automatically derive geometric parameters from a large number of DXA scans. In contrast to previous HSA studies, we were able to use this method to derive DFH measurements. FNW, DFH and HAL were all larger in males in contrast to females which is consistent with previous studies showing body measures are on average greater in males than females (31, 32). The geometric parameters obtained in this study compared favourably to those obtained by HSA in terms of correlation, but this study showed smaller mean measures likely due to differences in calculating the geometric parameters (12). For example, this study calculates FNW as the shortest distance across the femoral neck whereas HSA derives the FNW from its area measure making direct comparison difficult. Also, the HAL definition in this study does not include the pelvis unlike HSA; a previous study found a mean HSA-derived HAL of 104.7 mm, slightly higher than the mean in this study (96.7 mm) (33). Comparing geometric parameters derived in this study with other studies provides further face validity for our methods; mean FNW in this study is similar to that reported in the female only Study of Osteoporotic Fractures (mean FNW 29.0 mm v 30.7 mm) (7). DXA-derived DFH was very similar to DFH measured on proximal femurs removed during hemiarthroplasty in a previous cross-sectional study (mean 45.9 mm vs 44.9 mm respectively) (34).

The strengths of this study include its large sample size obtained through novel automated measures which have facilitated the description of DXA-derived hip geometry in UKB, including the derivation of DFH which is not provided by existing HSA software. A sample size of this magnitude paves the way for future use of genetic epidemiological methods, which can add further understanding of aetiology of hip shape and its role in the pathogenesis of HOA (26, 35). In addition, the use of DXA scans which require lower radiation exposure than X-rays makes them more desirable for screening (10, 36).

There are several limitations to this study. Firstly, this was an observational study, so we cannot infer causality. The UKB database contains predominantly Caucasian participants, limiting generalisability of findings to other populations and warranting further research in more ethnically diverse settings. In addition, the clinical outcomes examined (HESOA and THR) were not side specific and this may have biased our effect sizes towards the null. It was not possible to calculate neck-shaft angle or femoral shaft width as the outline points did not extend distally below the lesser trochanter. That said, prior studies suggested that the three parameters which were included may have the greatest relevance for hip OA (6, 7, 23). Finally, although HSA is an established methodology for deriving hip geometry, we were keen to explore the role of DFH, which is not calculated by HSA. Therefore, we developed a bespoke platform for deriving FNW, HAL and DFH from 85 points outlining the proximal femur and acetabulum. Reassuringly, FNW and HAL generated by both methods correlated closely in a subset of participants.

In conclusion, three geometric parameters (FNW, DFH and HAL) were automatically derived from high resolution DXA scans in UKB. Incorporating their inter-relatedness into regression models, strong independent associations were observed between FNW and HOA outcomes (rHOA, HESOA and THR). Weaker independent associations were seen between DFH and HAL, and rHOA. This work paves the way for clinical translation as it suggests geometric parameters derived from DXA are associated with HOA, including the risk of future hip replacement, but further work is needed to understand how to combine measures of hip morphology to better predict HOA risk and progression.

## Data Availability

All geometric parameters included in this study have been returned to UK Biobank and will be available at a forthcoming data release. Users must be registered with UK Biobank to access their resources [https://bbams.ndph.ox.ac.uk/ams/].

## Acknowledgements

This work has been conducted using the UK Biobank resource, access application 17295.

## Author contributions

All authors have made significant contributions to the conception and design of this study, the acquisition of data, its analysis and interpretation. All authors helped draft the article before approving the final version of this manuscript. Sophie Heppenstall takes responsibility for the integrity of the work in its entirety.

