## Supplementary Material for "Hip geometric parameters are associated with radiographic and clinical hip osteoarthritis: findings from a cross-sectional study in UK Biobank"

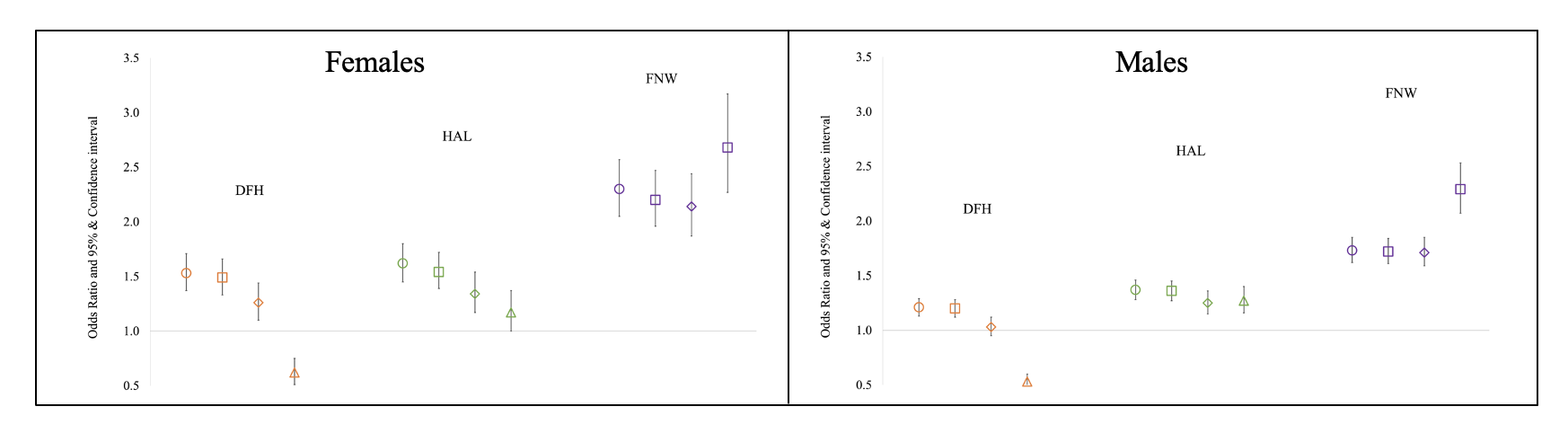
**Supplementary Material**

Supplementary Figure 1: Logistic regression results for the associations between geometric parameters – diameter of the femoral head (DFH), hip axis length (HAL) & femoral neck width (FNW) and radiographic hip osteoarthritis grade ≥ 2 in sex stratified analyses. Odds ratios with 95% CIs were plotted either side of the points. Circle symbol represents unadjusted analyses (model 1), square indicates adjustment for age and sex (model 2), diamond for age, sex, height and weight (model 3) and triangle for age, sex, height, weight, and remaining geometric parameters (model 4)*.*

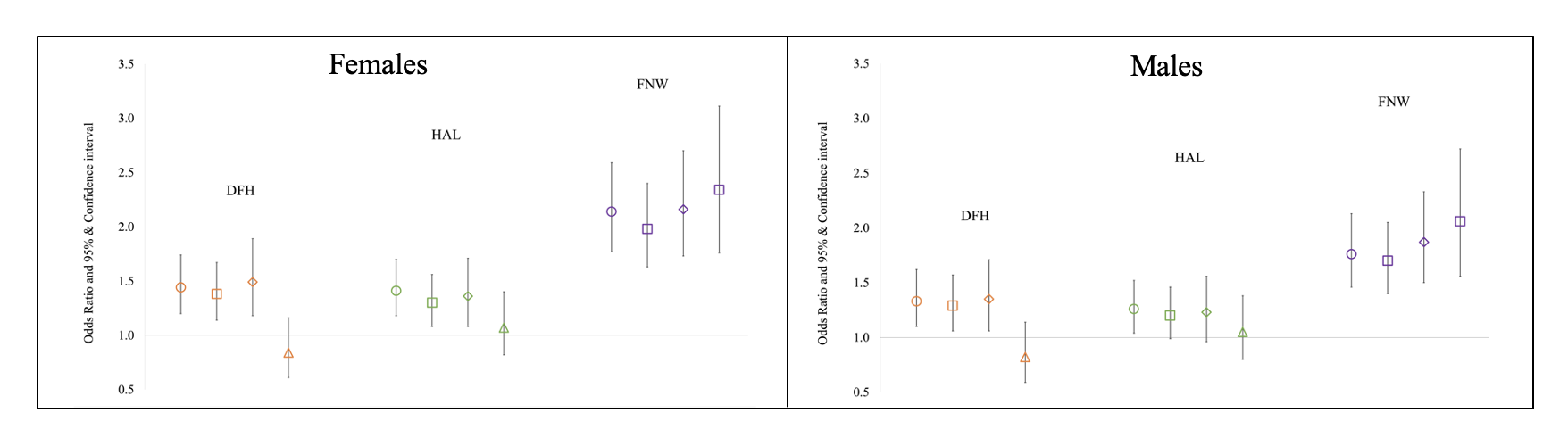
Supplementary Figure 2: Logistic regression results for the associations between geometric parameters – diameter of the femoral head (DFH), hip axis length (HAL) & femoral neck width (FNW) and hospital diagnosed osteoarthritis (HESOA) in sex stratified analyses. Odds ratios with 95% CIs were plotted either side of the points. Circle symbol represents unadjusted analyses (model 1), square indicates adjustment for age and sex (model 2), diamond for age, sex, height and weight (model 3) and triangle for age, sex, height, weight, and remaining geometric parameters (model 4)*.*

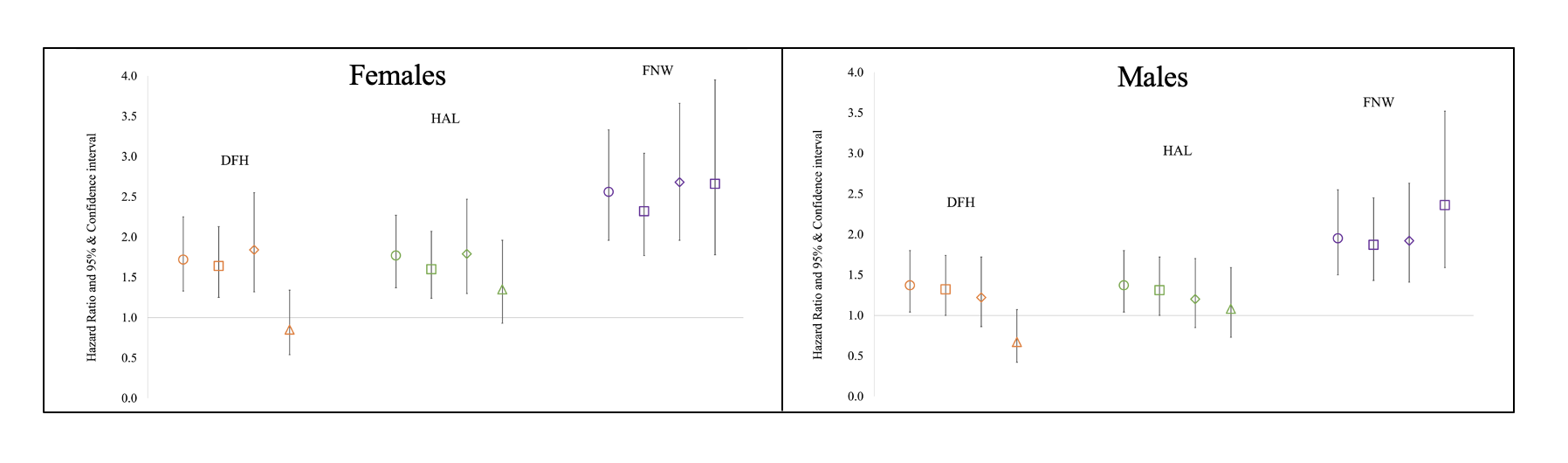
Supplementary Figure 3: Cox regression results for the associations between geometric parameters – diameter of the femoral head (DFH), hip axis length (HAL) & femoral neck width (FNW) and total hip replacement (THR) in sex stratified analyses. Hazard ratios with 95% CIs were plotted either side of the points. Circle symbol represents unadjusted analyses (model 1), square indicates adjustment for age and sex (model 2), diamond for age, sex, height and weight (model 3) and triangle for age, sex, height, weight, and remaining geometric parameters (model 4)*.*

Supplementary Table 1: Glossary of terms

| DFH | Diameter of the femoral head |
| --- | --- |
| DXA | Dual-energy x-ray absorptiometry |
| FNW | Femoral neck width |
| HAL | Hip axis length |
| HES | Hospital episode statistics |
| HESOA | Hospital diagnosed osteoarthritis |
| HOA | Hip osteoarthritis |
| HSA | Hip structural analysis |
| OA | Osteoarthritis |
| SSM | Statistical Shape Modelling |
| THR | Total Hip Replacement |
| UKB | UK Biobank |
| rHOA | Radiographic hip osteoarthritis |

Supplementary Table 2: Comparison between automatically derived GPs and by hip structural analysis.

| Geometric Parameter | **N** | **Hip Structural Analysis** | **Automated Method** | **Correlation R^2^** |
| --- | --- | --- | --- | --- |
|  |  | **Mean [Range]** | **Mean [Range]** |  |
| Femoral Neck Width /Narrowest neck width (NNW) (mm) | 1744 | 35.0 [27.3-49.2] | 31.9 [24.4-42.0] | 0.97 |
| Hip Axis Length (HAL) (mm) | 1689 | 115.1 [91.0-160.9] | 97.0 [77.5-127.3] | 0.93 |

Supplementary Table 3: Logistic regression/ Cox proportional hazard modelling results showing the association between geometric parameters and HOA outcomes in partially adjusted models.

|  | | **Grade ≥ 2 rHOA** | | **Grade ≥ 3 rHOA** | | **Grade 4 rHOA** | | **HESOA** | | **THR** | |
| --- | --- | --- | --- | --- | --- | --- | --- | --- | --- | --- | --- |
|  |  | **OR [95% CI]** | **p** | **OR [95% CI]** | **p** | **OR [95% CI]** | **p** | **OR [95% CI]** | **p** | **HR [95% CI]** | **p** |
| Model 1 | FNW | 1.85 [1.78-1.92] | 8.00 x 10^-223^ | 2.40 [2.22-2.59] | 1.00x10^-108^ | 2.86 [2.43-3.37] | 4.86x10^-36^ | 1.18 [1.09-1.29] | 1.30x10^-4^ | 1.23 [1.09-1.39] | 6.56x10^-4^ |
| Model 2 | FNW | 1.83 [1.73-1.94] | 2.50 x 10^-94^ | 2.92 [2.62-3.27] | 5.00x10^-80^ | 4.13 [3.31-5.16] | 6.03x10^-36^ | 1.83 [1.60-2.10] | 2.10x10^-18^ | 2.09 [1.73-2.52] | 3.03x10^-14^ |
| Model 1 | HAL | 1.66 [1.60-1.72] | 3.00x10^-154^ | 1.82 [1.69-1.96] | 2.80x10^-55^ | 1.89 [1.62-2.21] | 1.47x10^-15^ | 1.03 [0.94-1.12] | 0.55 | 1.08 [0.96-1.22] | 0.2 |
| Model 2 | HAL | 1.41 [1.33-1.49] | 7.83x10^-33^ | 1.59 [1.42-1.77] | 2.22x10^-16^ | 1.76 [1.40-2.21] | 1.33x10^-6^ | 1.25 [1.10-1.43] | 9.17x10^-4^ | 1.46 [1.21-1.76] | 7.71x10^-5^ |
| Model 1 | DFH | 1.58 [1.52-1.64] | 4.00x10^-128^ | 1.77 [1.64-1.90] | 2.40x10^-50^ | 1.90 [1.63-2.22] | 8.45x10^-16^ | 1.04 [0.95-1.13] | 0.42 | 1.06 [0.94-1.20] | 0.31 |
| Model 2 | DFH | 1.27 [1.20-1.34] | 6.20x10^-16^ | 1.51 [1.35-1.69] | 1.13x10^-12^ | 1.82 [1.45-2.30] | 3.73x10^-7^ | 1.33 [1.16-1.53] | 3.35x10^-5^ | 1.47 [1.22-1.78] | 7.30x10^-5^ |

Table shows logistic regression results for the associations between geometric parameters and radiographic hip osteoarthritis (rHOA) ≥ grade 2, ≥ grade 3, grade 4 and hospital diagnosed hip osteoarthritis (HESOA) and cox proportional hazard modelling results between geometric parameters and total hip replacement (THR) in unadjusted and partially adjusted combined sex analyses (models 1 & 2). Model 1 is the unadjusted model and model 2 included the covariates age and sex.

Supplementary Table 4: Logistic regression/ Cox proportional hazard modelling results showing the association between geometric parameters

| **Femoral Neck Width (FNW) (mm)** | **Grade ≥2 rHOA** | | **Grade ≥3 rHOA** | | **Grade≥ 4 rHOA** | | **Hospital diagnosis of HOA** | | **THR** | |
| --- | --- | --- | --- | --- | --- | --- | --- | --- | --- | --- |
|  | **OR [95% CI]** | **p** | **OR [95% CI]** | **p** | **OR [95% CI]** | **p** | **OR [95% CI]** | **p** | **HR [95%]** | **p** |
| Unadjusted (Model 1) | 2.3 [2.05-2.57] | 2.70x10^-47^ | 4.04 [3.19-5.10] | 1.59x10^-31^ | 6.70 [4.21-10.69] | 1.27x10^-15^ | 2.14 [1.77-2.59] | 5.27x10^-15^ | 2.56 [1.96-3.33] | 3.38x10^-12^ |
| Partially adjusted (Model 2) | 2.20 [1.96-2.47] | 5.20x10^-42^ | 3.73 [2.94-4.73] | 2.67x10^-27^ | 6.20 [3.86-9.98] | 5.44x10^-14^ | 1.98 [1.63-2.40] | 4.23x10^-12^ | 2.32 [1.77-3.04] | 8.36x10^-10^ |
| Partially adjusted (Model 3) | 2.14 [1.87-2.44] | 7.47x10^-30^ | 4.47 [3.39-5.90] | 2.18x10^-26^ | 7.83 [4.48-13.67] | 5.03x10^-13^ | 2.16 [1.73-2.70] | 1.38x10^-11^ | 2.68 [1.96-3.66] | 5.99x10^-10^ |
| Fully adjusted (Model 4) | 2.68 [2.27-3.17] | 9.70x10^-31^ | 6.08 [4.26-8.69] | 3.59x10^-23^ | 8.12 [3.95-16.70] | 1.28x10^-8^ | 2.34 [1.76-3.11] | 4.85x10^-9^ | 2.66 [1.78-3.95] | 1.48x10^-6^ |
| **Hip Axis Length (HAL) (mm)** | | | | | | | | | | |
| Unadjusted (Model 1) | 1.62 [1.45-1.80] | 7.63x10^-19^ | 2.30 [1.84-2.87] | 1.76x10^-13^ | 3.39 [2.19-5.26] | 4.44x10^-8^ | 1.41 [1.18-1.70] | 2.08x10^-4^ | 1.77 [1.37-2.27] | 1.04x10^-5^ |
| Partially adjusted (Model 2) | 1.54 [1.39-1.72] | 2.22x10^-15^ | 2.10 [1.67-2.63] | 1.24x10^-10^ | 3.08 [1.97-4.80] | 7.77x10^-7^ | 1.30 [1.08-1.56] | 5.57x10^-3^ | 1.60 [1.24-2.07] | 3.32x10^-4^ |
| Partially adjusted (Model 3) | 1.34 [1.17-1.54] | 1.99x10^-5^ | 2.27 [1.71-3.00] | 1.00x10^-8^ | 3.53 [2.03-6.14] | 8.35x10^-6^ | 1.36 [1.08-1.71] | 8.47x10^-4^ | 1.79 [1.30-2.47] | 3.75x10^-4^ |
| Fully adjusted (Model 4) | 1.17 [1.00-1.37] | 0.05 | 1.76 [1.27-2.44] | 7.09x10^-4^ | 2.05 [1.08-3.89] | 0.03 | 1.07 [0.82-1.40] | 0.61 | 1.35 [0.93-1.96] | 0.11 |
| **Diameter of the femoral head (DFH) (mm)** | | | | | | | | | | |
| Unadjusted (Model 1) | 1.53 [1.37-1.71] | 6.18x10^-14^ | 1.93 [1.52-2.44] | 4.41x10^-8^ | 3.26 [2.04-5.21] | 8.20x10^-7^ | 1.44 [1.20-1.74] | 1.40x10^-4^ | 1.72 [1.33-2.25] | 1.04x10^-5^ |
| Partially adjusted (Model 2) | 1.49 [1.33-1.66] | 2.16x10^-12^ | 1.83 [1.44-2.32] | 5.89x10^-7^ | 3.10 [1.93-4.98] | 4.04x10^-6^ | 1.38 [1.14-1.67] | 8.68x10^-4^ | 1.64 [1.25-2.13] | 3.32x10^-4^ |
| Partially adjusted (Model 3) | 1.26 [1.10-1.44] | 1.15x10^-3^ | 1.82 [1.35-2.43] | 6.65x10^-5^ | 3.46 [1.93-6.19] | 3.02x10^-5^ | 1.49 [1.18-1.89] | 8.47x10^-4^ | 1.84 [1.32-2.55] | 3.75x10^-4^ |
| Fully adjusted (Model 4) | 0.62 [0.51-0.75] | 6.37x10^-7^ | 0.42 [0.28-0.64] | 3.98x10^-5^ | 0.62 [0.27-1.42] | 0.26 | 0.84 [0.61-1.16] | 0.28 | 0.85 [0.54-1.34] | 0.11 |

and HOA outcomes in females.

Table shows logistic regression results for the associations between geometric parameters and radiographic hip osteoarthritis (rHOA) ≥ grade 2, ≥ grade 3, grade 4 and hospital diagnosed hip osteoarthritis (HESOA) and cox proportional hazard modelling results between geometric parameters and total hip replacement (THR) in unadjusted, partially and fully adjusted analyses in females (models 1 - 4). Model 1 is the unadjusted model, model 2 included the covariates age and sex, model 3 included model 2 plus height and weight and model 4 is the fully adjusted model including model 3 plus the remaining geometric parameters.

Supplementary Table 5: Logistic regression/ Cox proportional hazard modelling results showing the association between geometric parameters

| **Femoral Neck Width (FNW) (mm)** | **Grade ≥2 rHOA** | | **Grade ≥3 rHOA** | | **Grade 4 rHOA** | | **Hospital diagnosis of HOA** | | **THR** | |
| --- | --- | --- | --- | --- | --- | --- | --- | --- | --- | --- |
|  | **OR [95% CI]** | **p** | **OR [95% CI]** | **p** | **OR [95% CI]** | **p** | **OR [95% CI]** | **p** | **HR [95%]** | **p** |
| Unadjusted (Model 1) | 1.73 [1.62-1.85] | 4.10x10^-49^ | 2.74 [2.42-3.1] | 6.70x10^-57^ | 3.76 [2.93-4.83] | 1.95x10^-25^ | 1.76 [1.46-2.13] | 3.03x10^-9^ | 1.95 [1.50-2.55] | 7.60x10^-7^ |
| Partially adjusted (Model 2) | 1.72 [1.61-1.84] | 2.10x10^-57^ | 2.72 [2.40-3.09] | 9.00x10^-56^ | 3.69 [2.87-4.74] | 2.10x10^-24^ | 1.70 [1.40-2.05] | 5.02x10^-8^ | 1.87 [1.43-2.45] | 4.46x10^-6^ |
| Partially adjusted (Model 3) | 1.71 [1.59-1.85] | 5.10x10^-43^ | 3.29 [2.85-3.80] | 2.00x10^-58^ | 4.55 [3.41-6.08] | 1.00x10^-24^ | 1.87 [1.50-2.33] | 2.71x10^-8^ | 1.92 [1.41-2.63] | 4.14x10^-5^ |
| Fully adjusted (Model 4) | 2.29 [2.07-2.53] | 1.70x10^-27^ | 5.00 [4.15-6.04] | 9.20x10^-55^ | 8.61[5.90-12.57] | 6.56x10^-29^ | 2.06 [1.56-2.72] | 4.24x10^-7^ | 2.36 [1.59-3.52] | 2.33x10^-5^ |
| **Hip Axis Length (HAL) (mm)** | | | | | | | | | | |
| Unadjusted (Model 1) | 1.37 [1.28-1.46] | 6.03x10^-21^ | 1.47 [1.30-1.67] | 1.78x10^-9^ | 1.51 [1.16-1.96] | 2.17x10^-3^ | 1.26 [1.04-1.52] | 0.02 | 1.37 [1.04-1.80] | 0.02 |
| Partially adjusted (Model 2) | 1.36 [1.27-1.45] | 8.37x10^-20^ | 1.45 [1.28-1.65] | 7.02x10^-9^ | 1.46 [1.12-1.90] | 5.37x10^-3^ | 1.20 [0.99-1.46] | 0.06 | 1.31 [1.00-1.72] | 0.05 |
| Partially adjusted (Model 3) | 1.25 [1.15-1.36] | 9.50x10^-8^ | 1.47 [1.25-1.72] | 2.31x10^-6^ | 1.28 [0.91- 1.79] | 0.15 | 1.23 [0.96-1.56] | 0.10 | 1.20 [0.85-1.70] | 0.30 |
| Fully adjusted (Model 4) | 1.27 [1.16-1.40] | 6.70x10^-7^ | 1.27 [1.06-1.52] | 9.78x10^-3^ | 1.08 [0.74-1.57] | 0.69 | 1.05 [0.80-1.38] | 0.74 | 1.08 [0.73-1.59] | 0.71 |
| **Diameter of the femoral head (DFH) (mm)** | | | | | | | | | | |
| Unadjusted (Model 1) | 1.21 [1.13-1.29] | 3.70x10^-8^ | 1.44 [1.27-1.64] | 2.02x10^-8^ | 1.60 [1.23-2.08] | 5.05x10^-4^ | 1.33 [1.10-1.62] | 3.43x10^-3^ | 1.37 [1.04-1.80] | 0.02 |
| Partially adjusted (Model 2) | 1.20 [1.12-1.28] | 1.37x10^-7^ | 1.43 [1.25-1.62] | 5.57x10^-8^ | 1.56 [1.19-2.03] | 1.12x10^-3^ | 1.29 [1.06-1.57] | 0.01 | 1.32 [1.00-1.74] | 0.05 |
| Partially adjusted (Model 3) | 1.03 [0.95-1.12] | 0.46 | 1.41 [1.21-1.65] | 1.73x10^-5^ | 1.41 [1.02-1.96] | 0.04 | 1.35 [1.06-1.71] | 0.01 | 1.22 [0.86-1.72] | 0.26 |
| Fully adjusted (Model 4) | 0.53 [0.47-0.60] | 1.22x10^-26^ | 0.43 [0.34-0.54] | 1.46x10^-13^ | 0.31 [0.19-0.50] | 1.06x10^-6^ | 0.82 [0.59-1.14] | 0.24 | 0.67 [0.42-1.07] | 0.09 |

and HOA outcomes in males.

Table shows logistic regression results for the associations between geometric parameters and radiographic hip osteoarthritis (rHOA) ≥ grade 2, ≥ grade 3, grade 4 and hospital diagnosed hip osteoarthritis (HESOA) and cox proportional hazard modelling results between geometric parameters and total hip replacement (THR) in unadjusted, partially and fully adjusted analyses in males (models 1 - 4). Model 1 is the unadjusted model, model 2 included the covariates age and sex, model 3 included model 2 plus height and weight and model 4 is the fully adjusted model including model 3 plus the remaining geometric parameters.

Supplementary Table 6: Logistic regression results showing the association between the ratio of DFH to FNW and HOA outcomes in combined sex analyses.

|  | **Grade ≥ 2 rHOA** | | **Grade ≥ 3 rHOA** | | **Grade 4 rHOA** | | **HESOA** | | **THR** | |
| --- | --- | --- | --- | --- | --- | --- | --- | --- | --- | --- |
|  | **OR [95% CI]** | **p** | **OR [95% CI]** | **p** | **OR [95% CI]** | **p** | **OR [95% CI]** | **p** | **HR [95% CI]** | **p** |
| Unadjusted (Model 1) | 0.56 [0.54-0.58] | 2.00x10^-172^ | 0.38 [0.35-0.41] | 1.00x10^-109^ | 0.30 [0.25-0.36] | 7.70x10^-39^ | 0.74 [0.68-0.81] | 4.50x10^-11^ | 0.71 [0.63-0.81] | 1.50x10^-7^ |
| Partially adjusted (Model 2) | 0.64 [0.61-0.67] | 3.50x10^-88^ | 0.42 [0.38-0.46] | 1.60x10^-76^ | 0.32 [0.26-0.39] | 1.40x10^-30^ | 0.68 [0.62-0.75] | 1.80x10^-14^ | 0.65 [0.56-0.74] | 1.40x10^-9^ |
| Partially adjusted (Model 3) | 0.65 [0.62-0.68] | 2.80x10^-82^ | 0.42 [0.38-0.46] | 2.50x10^-74^ | 0.32 [0.26-0.39] | 3.10x10^-30^ | 0.70 [0.63-0.77] | 2.70x10^-12^ | 0.66 [0.57-0.76] | 1.10x10^-8^ |
| Fully adjusted (Model 4) | 0.64 [0.61-0.67] | 6.40x10^-83^ | 0.41 [0.38-0.46] | 2.30x10^-75^ | 0.31 [0.26-0.38] | 1.30x10^-30^ | 0.70 [0.63-0.77] | 2.70x10^-12^ | 0.66 [0.57-0.76] | 1.10x10^-8^ |

Table shows logistic regression results for the associations between the ratio of DFH to FNW and grade ≥ 2,3 and 4 radiographic hip osteoarthritis (rHOA) and hospital diagnosed hip osteoarthritis (HESOA) and cox proportional hazard modelling results between geometric parameters and total hip replacement (THR) in unadjusted, partially and fully adjusted combined sex analyses (models 1 - 4). Model 1 is the unadjusted model, model 2 included the covariates age and sex, model 3 included model 2 plus height and weight and model 4 is the fully adjusted model including model 3 plus the remaining geometric parameters.
